# Characterizing the molecular impact of *KMT2D* variants on the epigenetic and transcriptional landscapes in Kabuki Syndrome

**DOI:** 10.1101/2022.10.25.22280882

**Authors:** Youngsook L Jung, Christina Hung, Jaejoon Choi, Eunjung A Lee, Olaf Bodamer

**Author notes:** Correspondence (O.B.).

## Abstract

Kabuki Syndrome (KS) is a rare, multisystem disorder with a variable clinical phenotype. The majority of KS is caused by dominant loss-of-function mutations in *KMT2D* (lysine methyltransferase 2D). KMT2D mediates chromatin accessibility by adding methyl groups to lysine residue 4 of histone 3, which plays a critical role in cell differentiation and homeostasis. The molecular underpinnings of KS remain elusive partly due to a lack of histone modification data from human samples. Consequently, we profiled and characterized alterations in histone modification and gene transcription in peripheral blood mononuclear cells (PBMCs) from 33 patients with *KMT2D* mutations and 36 unaffected healthy controls. Our analysis identified unique enhancer signatures in H3K4me1 and H3K4me2 in KS compared to controls. Reduced enhancer signals were present for promoter-distal sites of immune-related genes for which co-binding of PBMC-specific transcription factors was predicted; thirty-one percent of super-enhancers of normal blood cells overlapped with disrupted enhancers in KS, supporting an association of reduced enhancer activity of immune-related genes with immune deficiency phenotypes. In contrast, increased enhancer signals were observed for promoter-proximal regions of metabolic genes enriched with *EGR1* and *E2F2* motifs, whose transcriptional levels were significantly increased in KS. Additionally, we identified approximately 100 de novo enhancers in genes, such as in *MYO1F* and *AGAP2*. Together, our results underscore the effect of KMT2D haploinsufficiency on (dys)regulation of enhancer states and gene transcription and provide a framework for the identification of therapeutic targets and biomarkers in preparation for clinical trial readiness.

## INTRODUCTION

Kabuki Syndrome (KS) is a rare, heterogeneous, congenital malformation disorder that follows a broad and variable clinical spectrum characterized by a unique facial gestalt, mild to moderate intellectual disability, developmental delays, hearing loss, muscle hypotonia, and structural and/or functional anomalies of cardiac, endocrine, renal and orthopedic systems.(Niikawa et al. 1981; Kuroki et al. 1981; Adam et al. 2019) Abnormal regulation of terminal B-cell differentiation in KS leads to variable immune deficiency with low immunoglobulin (IgG and IgA) levels that may be associated with inadequate antibody response to vaccines and increased susceptibility to recurrent and potentially life-threatening infections.(Y. Li et al. 2011; Micale et al. 2011; Banka et al. 2012; Ming et al. 2005; Lindsley et al. 2016; Stagi et al. 2016; Adam, Hudgins, and Hannibal 2011) KS occurs in approximately 1:32,000 individuals and currently has no approved therapies, despite a significant disease burden.(Adam et al. 2019; Theodore-Oklota et al. 2022) Although it is known that KS is due to dominant loss-of-function mutations in autosomal *KMT2D* (lysine methyltransferase 2D, 80-90%, KS1, MIM: 147920) or X-linked *KDM6A* (lysine demethylase 6A, 5-8%, KS2, MIM: 300867) respectively,(Ng et al. 2010; Lederer et al. 2012) its underlying disease mechanisms are not fully elucidated. Most importantly, there is a limited understanding of the downstream pathways that are affected in KS and how they relate to the clinical phenotype spectrum.

Transcriptional regulation is a highly specialized and temporally orchestrated, tissue-specific process during early embryogenesis and development, which determines cell fate, cell cycle progression, stem cell function, and ultimately normal embryogenesis.(Fasciani et al. 2020) The complexity and importance of perfectly concerted transcriptional control necessitate a number of regulatory mechanisms, including chromatin and histone modifications. The addition or removal of histone marks, such as acetyl- and methyl-groups, changes the conformation of chromatin and renders genes accessible or inaccessible to transcription factor (TF) complexes.(Fasciani et al. 2020)

For gene activation, pioneer TFs, along with other cell-type-specific TFs, recruit KMT2D, a lysine histone methyl-transferase to prime enhancer regions through the addition of one methyl group (monomethylation) to the fourth lysine of histone 3 (H3K4), thereby promoting open chromatin.(Ng et al. 2010; Froimchuk, Jang, and Ge 2017) KMT2D associates with WRAD (WDR5, RbBP5, ASH2L, DPY30), KDM6A, NCOA6, PA1, and PTIP into one protein complex (ASCOM complex) which is critical for its H3K4 methyltransferase activity and stabilization of KDM6A (also known as UTX).(Froimchuk et al. 2017) Additionally, KMT2D facilitates the binding of the two H3K27 acetyltransferases CREB-binding protein (CBP) and p300 on enhancers, thereby promoting tissue-specific gene expression.

Loss of KMT2D function is therefore expected to alter chromatin states, in particular enhancer states, and to subsequently cause gene dysregulation. In addition to the association with KS, mutations in *KMT2D* have been described in association with various cancers. *C*onsidered a tumor suppressor gene, *KMT2D* is one of the most frequently mutated genes in cancer for reasons yet unknown.(J. Zhang et al. 2015; Ortega-Molina et al. 2015) In B-cell lymphoma and leukemia, reduced activity of KMT2D leads to tumor proliferation and disruption of tumor suppressor pathways, contrasting other cancers, including breast cancer, in which reduced expression of KMT2D causes decreased proliferation, suggesting diverging tissue-type-dependent effects.

There have been recent advances in understanding the specific role of KMT2D in the KS disease process.(Fahrner et al. 2019; Carosso et al. 2019; Huisman et al. 2021; Bjornsson et al. 2014) Previous studies largely focusing on human blood DNA methylation profiles showed that the DNA methylation profiles differ between KS and unaffected controls.(Aref-Eshghi et al. 2017, 2018; Sobreira et al. 2017) While differentially methylated regions may be useful as future biomarkers, they are of limited use for elucidating regulatory mechanisms in KS pathology as *KMT2D* mutations primarily affect histone modifications and only subsequently exert their effect on DNA methylation.

In this study, we concomitantly profiled genome-wide histone modification along with gene expression in peripheral blood from 33 KS individuals with deleterious *KMT2D* mutations to study the impact on chromatin states and gene regulation. By integrating epigenetic and transcriptomic data, we identified changes in histone modification, gene regulation and regulatory pathways in KS. Importantly, we found unique chromatin changes that may serve as future drug targets or biomarkers, preparing for KS for clinical trial readiness.

## MATERIAL AND METHODS

### Subjects

Individuals of any age with a molecularly confirmed and clinical diagnosis of KS, due to likely pathogenic or pathogenic variants in *KMT2D*, and age/gender-matched healthy controls were eligible to participate in this study following written informed consent. Individuals with KMT2D-related disorder due to missense variants in exons 38 and 39 of *KMT2D*,(Cuvertino et al. 2020) as well as individuals with KS affected by an additional genetic disorder, were excluded from participation in this study. All subjects were deidentified and family IDs are only known to the principle investigator (OB) and the immediate research staff. The study was approved by the Institutional Review Board of Boston Children’s Hospital (IRB-A00026691).

The molecular diagnosis of KS was confirmed by genetic testing through CLIA-certified clinical laboratories. Parental testing was conducted and showed de novo occurrence of KMT2D variants in the majority of cases. The interpretation of variant classification reflects the classification reported by the diagnostic laboratory and follows the guidelines of The American College of Medical Genetics and Genomics and the Association for Molecular Pathology.(Richards et al. 2015) The variant types were annotated and the genomic positions were extracted using the Mutalyzer tool.(Lefter et al. 2021)

### Sample preparation, library construction and sequencing for ChIP-seq data

PBMCs were isolated from NaHep samples using the Lymphoprep™/SepMate™ system (StemCell Technologies) according to the manufacturer’s specifications and cryopreserved in 10% DMSO v/v FBS. For cell fixation, cells were thawed, counted and 6 million cells per sample were fixed in a 1% formaldehyde solution for 15 min at room temperature. The fixation was stopped using 0.125M Glycine for 5 min and cell pellets were washed in 0.5% Igepal in PBS and 1mM PMSF in 0.5% Igepal-PBS. Snap frozen cell pellets were then shipped on dry ice to ActiveMotif for chromatin extraction and Histone Path™ ChIP-Seq experiments. Sequences were generated on the illumina NextSeq 500 platform. Takeda Pharmaceutical, Cambridge, MA, provided financial support for the ChIP-Seq experiments. Raw data was returned for further bioinformatics analyses as described in the following sections.

### Sample preparation, RNA-extraction, library construction and sequencing for RNA-seq data

Total RNA was extracted from whole blood following the manufacturer’s protocol for PAXgene Blood RNA Kit (Qiagen). Following the assessment of RNA concentration and quality (RIN>7.2) by Agilent 2100 or Fragment Analyzer, the illumina TruSeq Stranded Total RNA with Ribo-Zero Globin library preparation was used, which includes ribosomal RNA and globin mRNA removal. A minimum of 10Gb of 100bp paired-end reads were generated per sample. All library preparations and sequencing was conducted by BGI Americas on the illumina HiSeq 4000 platform. Raw data was returned for further bioinformatics analysis as described in the following sections.

### Data processing and analysis of ChIP-seq data

For ChIP-seq data, the reads were mapped to the GRCh38 human reference sequence using BWA (Burrows-Wheeler Aligner) with default parameters.(H. Li and Durbin 2009) Only uniquely mappable reads were used for the downstream analysis. Significantly enriched regions were detected and normalized IP (immunoprecipitation) enrichment profiles were generated using MACS2 (Model-bases analysis of ChIP-seq 2) call peak function with default parameters.(Feng et al. 2012) To assess the data quality, the FRiP (fraction of reads in peaks) scores were calculated and correlations were measured between samples. The samples with low FRiP scores or correlation efficiencies were excluded from the downstream analysis.

For the principal component analysis (PCA), peaks from each individual (both KS and controls) were used to identify the union peak regions. Among the union peak regions, the peaks showing consistent signal intensities across both cohorts (KS vs control) were excluded after calculating a coefficient of variation (standard deviation over mean). The PCA analysis was performed using the read counts in the remaining differing peak sets.

For the detection of differential peaks between groups (KS vs control), Diffbind R package was used for the union peak sets.(Ross-Innes et al. 2012; Gupta et al. 2007) For visualization of the differential peaks in a heatmap, values in each row were normalized. The differential peaks were divided into two groups--a group with increased and a group with decreased signals in KS compared to control. For both groups, the enriched gene ontology terms were identified using the Great tool with default parameters.(McLean et al. 2010) For motif analysis, the TSS (transcription start site)-proximal and TSS-distal regions were further defined. TSS information was obtained from the UCSC genome browser table for GRCh38. Peaks within a 2 kb distance from TSS were defined as TSS-proximal whereas the peaks > 2 kb from TSS were deemed TSS-distal. DNA sequences were extracted within a 100 bp window from peak summits and known motifs were matched using MEME TOMTOM.(Gupta et al. 2007) We obtained the motif sequences for the search from JASPAR (Khan et al. 2018) and Jolma et al in 2013.(Jolma et al. 2013) The genomic locations of TF binding were predicted using MEME FEMO(Bailey et al. 2009) and used for the pathway enrichment analysis for a subset of TFs. To associate increased H3K4me2 sites in KS with respect to EGR1 binding, we used the publicly available EGR1 ChIP-seq data set from GM12878. To associate changes in histone modifications observed in our KS cohort with publicly available DNA methylation changes in KS, DNA methylation data from individuals with KS and control were downloaded from Sobreira et al (Sobreira et al. 2017) (GSE116300) and reanalyzed after performing lift over from hg18 to hg38.

We identified super-enhancers based on the intensity and breadth of H3K27ac signal in GM12878 following the methods described in Whyte et al (Whyte et al. 2013) and determined the intersection of the top 2000 differential peaks of H3K4me1 and H3K4me2 in KS compared with control and the super-enhancer regions.

### Analysis of RNA-seq data

RNA sequence reads were mapped using STAR aligner with default parameters.(Dobin et al. 2013) The transcriptional reads for each annotated gene were estimated using Htseq.(Anders, Pyl, and Huber 2015) Differentially expressed genes between KS and controls were determined using DESeq2 after adjusting for sex, age and batch effect.(Love, Huber, and Anders 2014) To associate genes with peaks of H3K4me1/2/3, the closest gene was assigned for each peak. If the gene has multiple H3K4me1/2/3 peaks, the peak with the lowest p-value was assigned to the gene. The TF list was obtained from AnimalTFDB.(Hu et al. 2019)

To infer transcriptional regulators such as TFs or chromatin regulators underlying differential expressions in KS, we used the method by Qin et al (Qin et al. 2020) which integrates TF binding sites from public ChIP-seq data in different tissue and cell types with differentially expressed genes. Briefly, using ∼10,000 available public ChIP-seq data sets, the regulatory potential score of each TF is calculated for differentially expressed genes with the assumption that the effect of the TF binding on gene expression exponentially decays with distance and that the contribution of multiple binding sites is additive. We used differentially expressed genes in KS compared to control with q-value (or FDR, false discovery rate) = 0.1.

## RESULTS

### Summary of cohort, variants and datasets

To generate RNA-seq and ChIP-seq data, we collected peripheral blood samples from 33 KS individuals with mutations in *KMT2D* (females: 23, mean age: 9.4 years, range: 0.9-33.3 years) and from 36 healthy individuals as age or sex-matched controls (females: 22, mean age: 30.8 years, range: 4.9-64.8 years) from either healthy siblings (age-matched), parents (sex-matched) of KS individuals or unrelated age-matched healthy children (see details in **Table S1**).

The variants in *KMT2D* of our cohort are summarized in **Table 1** and **Figure 1A**. The majority of variants were truncating (60.6%) and located before the SET domain, suggesting a loss of function of *KMT2D. KMT2D* has 54 exons. Although the variants were found across the *KMT2D* gene on chromosome 12, the distribution in the genomic positions indicates potential mutation hotspots depending on variant types. Frameshift variants were more prevalent in exon 10, while splice site variants clustered in introns 37-38, nonsense mutations in exon 39 and missense mutations in exon 48 (**Figure 1B**). In particular, exons 39 and 48 were most frequently mutated (36%, 12/33), consistent with the prior studies. (Hannibal et al. 2011; Banka et al. 2012). A total of 8 variants including 6 nonsense mutations (more than half, 6/8) were identified in exon 39, which encodes long polyglutamine tracts, and 4 variants were found in exon 48. Interestingly, most missense mutations (5/6) were located in F/Y-rich domains (FYRN and FYRC), transcription activation domains involved in TF bindings.(Park et al. 2010)

**Table 1.** Features of subjects and mutations in *KMT2D*. ** indicates a twin.

| Subject | Sex | Age at enrollment | *KMT2D* variant (NM_003482.3) | Variant type | Genomic position in hg19 | Parental origin |
| --- | --- | --- | --- | --- | --- | --- |
| **S1** | male | 4-10 years | c.839+1del, intron 6 | splice site | 12:49447259 | n.d. |
| **S2** | female | 0-3 years | c.12844C>T (p.Arg4282*), exon 39 | nonsense | 12:49425644 | de novo |
| **S3** | male | 0-3 years | c.5104C>T (p.Arg1702*), exon 21 | nonsense | 12:49438067 | maternal |
| **S4** | male | 11-15 years | c.13450C>T (p.Arg4484*), exon 39 | nonsense | 12:49425038 | n.d. |
| **S5** | female | 16-30 years | c.6183+3G>T, intron 29 | splice site | 12:49435697 | n.d. |
| **S6** | female | 11-15 years | c.16437del (p.Asn5480Thrfs*7), exon 53 | frameshift | 12:49415910 | n.d. |
| **S7** | male | 16-30 years | c.9703_9704del (p.Lys3235Aspfs*15), exon 34 | frameshift | 12:49431435 | n.d. |
| **S8** | female | 11-15 years | c.10740+1G>A, intron 38 | splice site | 12:49427849 | n.d. |
| **S9** | female | 0-3 years | c.2657del (p.Pro886Leufs*44), exon 10 | frameshift | 12:49444813 | de novo |
| **S10** | female | 16-30 years | c.7481dup (p.Ala2496Serfs*10), exon 31 | frameshift | 12:49434073 | de novo |
| **S11** | female | 11-15 years | c.13689C>T (p.Pro4563=), exon 41 | likely to affect splicing | 12:49424534 | n.d. |
| **S12** | male | 0-3 years | c.15641G>T (p.Arg5214Leu), exon 48 | missense | 12:49420108 | de novo |
| **S13** | male | 11-15 years | c.16052G>A (p.Arg5351Gln), exon 50 | missense | 12:49418361 | n.d. |
| **S14** | female | 4-10 years | c.6086del (p.Pro2029Leufs*18), exon 28 | frameshift | 12:49435896 | n.d. |
| **S15** | male | 11-15 years | c.12395del (p.His4132Profs*10), exon 39 | frameshift | 12:49426093 | n.d. |
| **S16** | female | 4-10 years | c.13450C>T (p.Arg4484*), exon 39 | nonsense | 12:49425038 | n.d. |
| **S17** | female | 16-30 years | c.10507+2T>G, intron 37 | splice site | 12:49428191 | n.d. |
| **S18** | male | 4-10 years | c.12592C>T (p.Arg4198*), exon 39 | nonsense | 12:49425896 | de novo |
| **S19** | male | 4-10 years | c.2533del (p.Arg845Glyfs*85), exon 10 | frameshift | 12:49444938 | n.d. |
| **S20** | female | 0-3 years | c.10507+2T>G, intron 37 | splice site | 12:49428191 | n.d. |
| **S21** | female | 11-15 years | c.11263C>T (p.Gln3755*), exon 39 | nonsense | 12:49427225 | n.d. |
| **S22** | female | 16-30 years | c.10813C>T (p.Gln3605*), exon 39 | nonsense | 12:49427675 | de novo |
| **S23** | female | 0-3 years | c.6295C>T (p.Arg2099*), exon 31 | nonsense | 12:49435258 | de novo |
| **S24** | female | 31-50 years | c.16469_16472del (p.Lys5490Argfs*18), exon 53 | frameshift | 12:49415876_49415879 | de novo |
| **S25\*\*** | female | 4-10 years | c.15467A>G (p.Tyr5156Cys), exon 48 | missense | 12:49420282 | de novo |
| **S26\*\*** | female | 4-10 years | c.15467A>G (p.Tyr5156Cys), exon 48 | missense | 12:49420282 | de novo |
| **S27** | female | 0-3 years | c.5645-2A>G, intron 25 | splice site | 12:49436663 | maternal |
| **S28** | female | 16-30 years | c.15536G>A (p.Arg5179His), exon 48 | missense | 12:49420213 | n.d. |
| **S29** | female | 0-3 years | c.349C>T (p.Gln117*), exon 3 | nonsense | 12:49448362 | de novo |
| **S30** | female | 11-15 years | c.8859_8861delinsCA (p.Lys2953Asnfs*51), exon 34 | frameshift | 12:49432278_49432280 | n.d. |
| **S31** | male | 4-10 years | c.16295G>A (p.Arg5432Gln), exon 51 | missense | 12:49416416 | de novo |
| **S32** | male | 4-10 years | c.2713del (p.Glu905Asnfs*25), exon 10 | frameshift | 12:49444756 | de novo |
| **S33** | femal<br>e | 4-10 years | c.11386_11411del<br>(p.Gln3796Cysfs*207), exon 39 | frameshift | 12:494270<br>80_49427<br>105 | n.d. |

**Figure 1.**
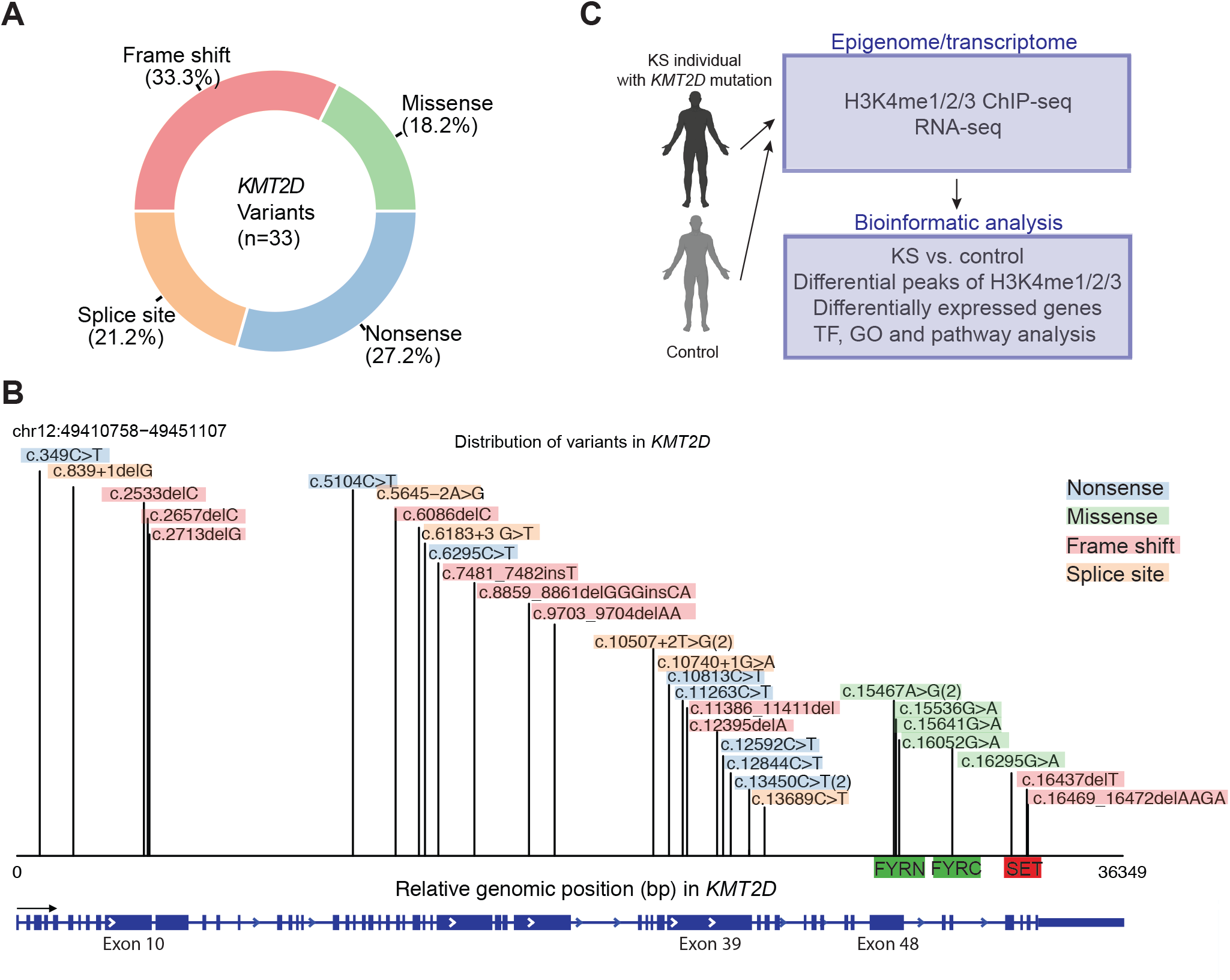
Summary of *KMT2D* mutations from KS individuals and datasets generated from this study. **A**. Proportions of *KMT2D* variant types from our KS cohort. Red, frame shift; blue, nonsense; green, missense; orange, splice site mutations. **B**. Variants and their mutation types across the *KMT2D* gene from our KS cohort. The genomic positions of each mutation site are shown along with the *KMT2D* gene where the transcription start direction is indicated with an arrow. The leftmost position indicates the transcription start site (TSS). (2) denotes mutations from two KS individuals. FYRN: F/Y-rich N-terminus domain, FYRC: F/Y-rich C-terminus domain. The same color codes are used as in A. **C**. Overview of this study design.

To characterize the alterations of chromatin and transcriptional states in KS and better understand the underlying regulatory mechanisms in KS, we generated histone modification profiles using ChIP-seq for H3K4me1, H3K4me2 and H3K4me3 which are known to be modified by KMT2D, along with RNA-seq data to study gene transcription. The overview of this study design is shown in **Figure 1C**.

### Distinctive enhancer signature in KS

We hypothesized that mutations in *KMT2D* will alter chromatin states and thereby affect the chromatin landscape in KS. H3K4me1 is found in primed enhancer regions that mark regulatory regions for enhancer activation in the later developmental stages or stimuli. These H3K4me1 marks will subsequently induce the enrichment for H3K27ac at the same enhancer regions. The H3K4me1 marks are typically located in promoter-distal sites, whereas H3K4me2, which is also associated with enhancer regions, enriched at both promoter-proximal regions as well as distal regions. In contrast, H3K4me3 is a marker for active promoters.

The principal component analysis for variable genomic regions among individuals revealed distinctive enhancer mark profiles of H3K4me1 (**Figure S1, Methods**) and H3K4me2 (**Figure 2A**), separating KS from control samples with greater distinction for H3K4me2. In contrast, H3K4me3 data could not be grouped according to the presence of KMT2D mutations. Rather, H3K4me3 data showed a tendency to cluster depending on genetic background (**Figure S1**). Similarly, transcriptional data (**Figure S1**) did not separate KS samples from controls. This suggests that *KMT2D* mutations affect enhancer states to a greater degree than promoter states, and that the enhancer signature of KS is different from healthy controls.

**Figure 2.**
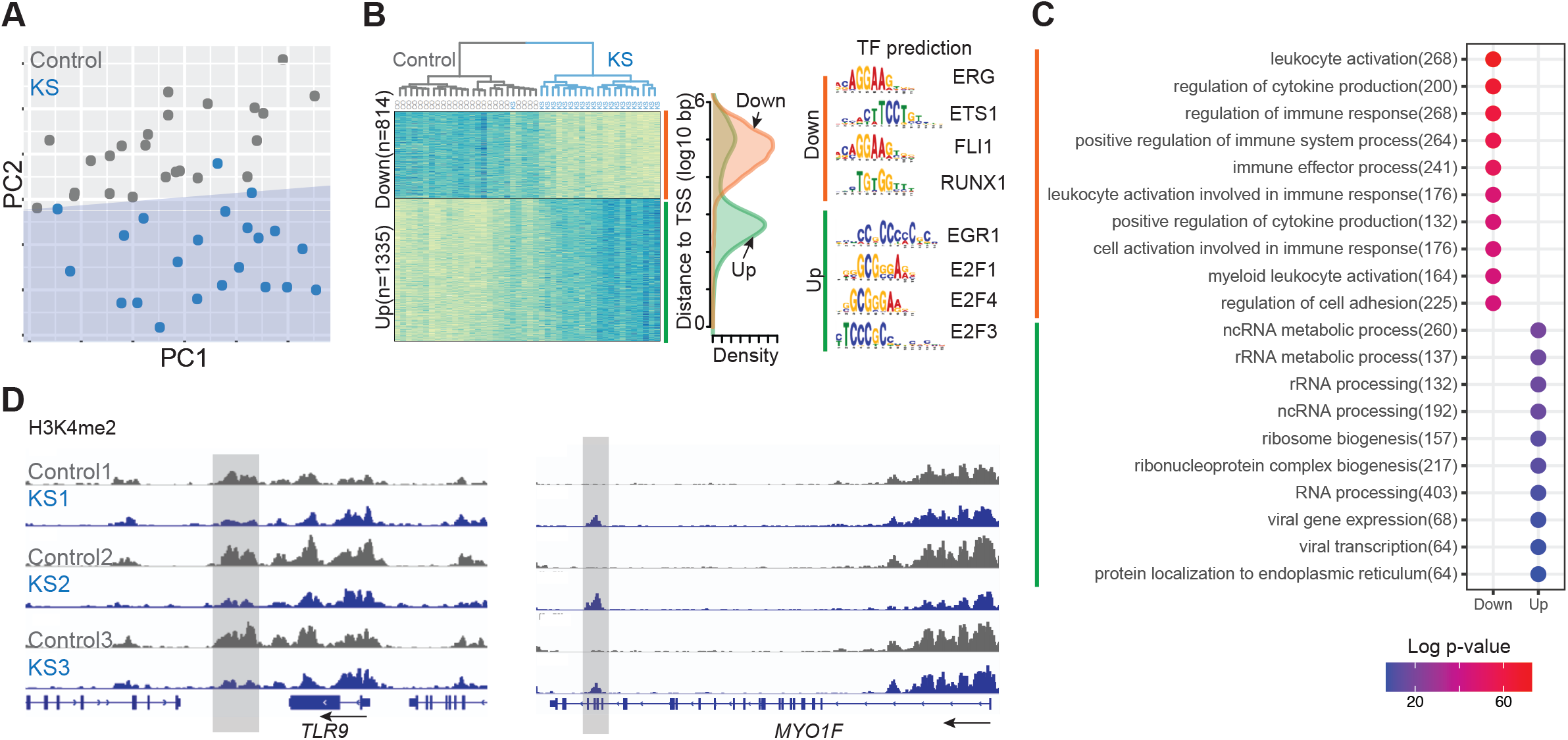
Alterations of enhancer state in KS individuals compared to controls. **A**. PCA analysis of variable H3K4me2 peaks for KS and control. Each dot represents an individual sample. Blue, KS; gray, control. **B**. Differential H3K4m2 peaks between KS and control. *<u>Left</u>*: row-normalized H3K4me2 signal heatmap of differential signals between KS and control. The hierarchical clustering results are shown on the top of the heatmap plot. Blue, high signal intensity; yellow, low signal intensity. *<u>Middle</u>*: distribution of distances of the differential peaks from TSSs. *<u>Right</u>*: top TF motifs predicted in differential H3K4me2 peaks between KS and control. Orange, decreased H3K4me2 peaks in KS compared to control; green, increased peaks in KS. **C**. Top gene ontology (GO) terms enriched for differential H3K4me2 peaks between KS and control. **D**. Representative of differential H3K4me2 sites between KS and control. Left: a representative of decreased H3K4me2 peaks in KS for *TLR9*. Blue, KS; gray, control. A gray box indicates significantly decreased H3K4me2 peaks in KS. An arrow indicates the transcriptional direction. Right: a representative of increased H3K4me2 peaks in KS for *MYO1F*. A gray box indicates significantly increased H3K4me2 peaks in KS.

### Regulatory program underlying altered enhancer signals in KS

We further characterized the alterations of enhancer marks H3K4me1 and H3K4me2 in KS compared to control. H3K4me2, an enhancer mark, was more distinctive between KS and control in PCA analysis. Hierarchical clustering results identified 2149 significantly changed H3K4me2 sites (814 decreased vs 1335 increased sites, q = 0.001, **Figure 2B, Methods**) confirming grouping according to genotypes, which is in alignment with the PCA result. Because loss of function of KMT2D is expected to cause a decrease of H3K4me2, a higher number of increased sites was unexpected but was consistent with a recent report in mouse B-cells.(Luperchio et al. 2021)

Furthermore, detailed examinations of H3K4me2 alterations revealed the association with two different target groups. Decreased sites were located at promoter-distal regions (**Figure 2B**) with enrichment of immune-related GO terms (**Figure 2C**) and were predicted to be bound by blood-cell-specific TFs through motif analysis. These blood-specific TFs, *ERG* (ETS Transcription Factor), *ETS1* (ETS Proto-Oncogene 1), *Fli1* (Friend Leukemia Integration 1) and *RUNX1* (Runt-Related Transcription Factor 1), play a crucial role in blood differentiation and development (**Figure 2B, Table S2**). A representative of significantly decreased H3K4me2 signals is shown in *TLR* (Toll-like receptor 9, top 4) where the decreased peak is located at the downstream intergenic region of the gene (**Figure 2D**). TLR plays an important role in the immune system and is implicated in the pathogenesis of autoimmune diseases.(Baccala et al. 2007) Similarly, we observed decreased H3K4me1 signals in promoter-distal sites of immune-related genes (**Figure S2**). In general, for the significantly changed H3K4me1 regions (q = 0.05), the changes of H3K4me1 signals in KS compared to control were correlated well with the changes of H3K4me2 signals (*r* = 0.92, changes of 99% peaks regions in the same direction, **Figure S2**).

While decreased H3K4me2 signals could be explained by the loss of function of *KMT2D*, the observed increases in H3K4me2 sites were unexpected. Therefore, we performed more in-depth analyses to explore underlying mechanisms for the increase in H3K4 dimethylation. Contrasting to the GO term associations of decreased H3K4me2 marks, the increased H3K4me2 peaks were enriched for metabolism-related GO terms and were found near promoter regions with the enrichment of the motifs for TFs associated with proliferation and inflammation such as the *EGR1* and *E2F* family (**Figure 2B, Table S2**). The expression changes of the TFs enriched in the increased H3K4me2 sites revealed that the expression levels of *EGR1* and *E2F2* increased by more than two-fold in KS patients compared to controls based on RNA-seq data (**Figure 3A**). In addition, differential expression of genes involved in cell cycles including *E2F3, E2F4* and *CCND1* were found to be increased, although the changes did not reach statistical significance.

**Figure 3.**
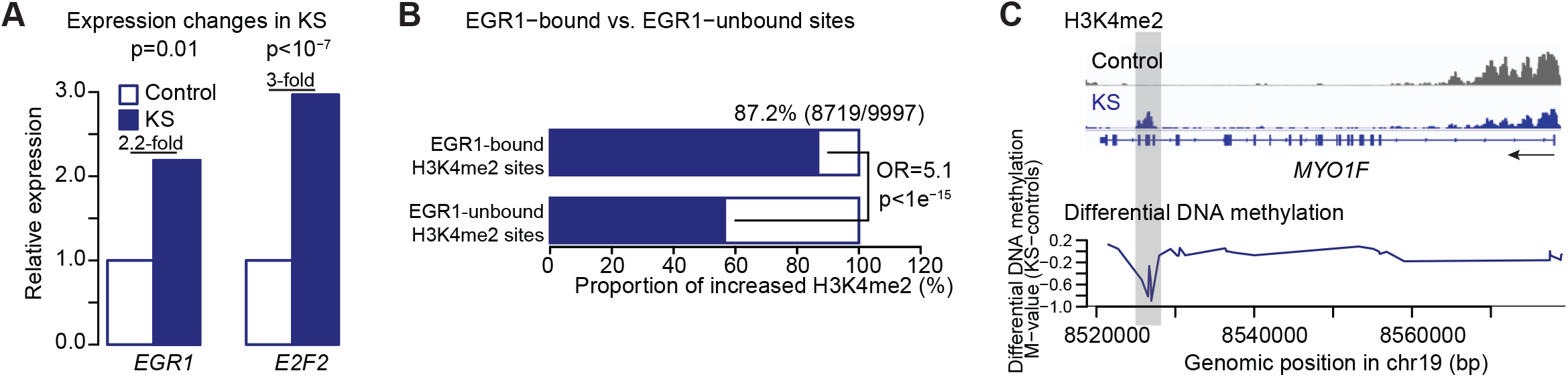
Increased H3K4me2 sites in KS compared to control. **A**. Expression changes of *EGR1* and *E2F2* in KS whose motifs were most enriched for increased H3K4me2 peaks in KS compared to control. The bar graph shows the expression levels of these genes significantly increased in KS. P-values from a negative binomial model. **B**. Fraction of the increased H3K4me2 peaks in KS for *EGR1* targets vs. non-*EGR1* targets. P-values by Fisher’s exact test. **C**. Differential DNA methylation profile of *MYO1F*. From the top, H3K4me2 profiles from KS individual and its control, and differential DNA methylation profile between KS and control, respectively. An arrow indicates the transcriptional direction. A gray box indicates the genomic regions overlapping with significantly increased H3K4me2 peaks and hypomethylated regions in KS compared to control.

We next examined the relationship between *EGR1* binding and H3K4me2 changes in KS using publicly available *EGR1* ChIP-seq data profiled in blood cells (**Methods**). The proportion of increased H3K4me2 sites at the promoter-proximal regions was drastically different between *EGR1*-bound sites and *EGR1*-unbound sites (**Figure 3B**). Among the *EGR1*-bound H3K4me2 sites, about 90% were increased in H3K4me2 signals, whereas among the H3K4me2 sites not targeted by EGR1, only half (about random expectation) showed increased H3K4me2 in KS, which is statistically significant (Odds ratio > 5 and *p*-value < 10^−15^ by Fisher’s exact test). Furthermore, the EGR1 target genes with the increased H3K4me2 showed a significant increase in expression (**Figure S2**) and were enriched in the pathways associated with hypoxia, cell cycles, Notch signaling and transcriptional regulation by the RUNX family (**Figure S2**). Altogether, these results suggest that KMT2D haploinsufficiency results in 1) reduced enhancer activity of immune-related genes and 2) overexpression of *EGR1* and *E2F2* associated with more open chromatin states, leading to the upregulation of target genes.

### De novo enhancer sites in KS individuals

Among the significantly increased H3K4me2 sites, we identified 135 de novo H3K4me2 peaks in KS (**Table S3**). One of the examples of de novo H3K4me2 peaks in KS is located in the intragenic region of *MYO1F* (Myosin 1F) (**Figure 2D**, top increased H3K4me2 site in KS compared to control). Several prior studies reported *MYO1F* as one of the most differentially hypomethylated regions in KS samples compared to controls. (Aref-Eshghi et al. 2017; Butcher et al. 2017; Aref-Eshghi et al. 2018; Sobreira et al. 2017) Comparisons with the previously published DNA methylation profile confirmed that the de novo H3K4me2 peak precisely overlapped with the hypomethylation region in KS (**Figure 3C**). Another example of de novo H3K4me2 sites is the gene *AGAP2* (ArfGAP With GTPase Domain 2, **Figure S3**). Similarly, several previous studies comparing blood DNA methylation profiles between KS and controls reported *AGAP2* as one of the hypomethylated regions in KS. The de novo H3K4me2 peak in *AGAP2* overlapped with the hypomethylated region in KS. In general, de novo H3K4me2 peaks in KS were less methylated in KS compared to controls based on available DNA methylation data (**Figure S3**). These histone and DNA methylation patterns can potentially serve as biomarkers for KS for clinical trial readiness.

### Association with super-enhancers

KMT2D deposits H3K4me1 and H3K4me2 marks at genomic targets, followed by p300 interaction mediated through the ASCOM complex, along with co-binding of TFs. This leads to H3K27ac and a fully active enhancer state. (Xu et al. 2018) The loss of function of KMT2D is expected to alter chromatin states in active enhancers as well as super-enhancers, which is supported by a previous study.(Alam et al. 2020) Super-enhancers are a set of enhancers typically identified by the strong and broad signals of H3K27ac and are known to play a critical role in development, cell identity and disease.(Hnisz et al. 2013; Lovén et al. 2013; Whyte et al. 2013) In our study, we found that approximately 31% of super-enhancers in lymphoblast cells (**Figure 4A**), which are defined by H3K27ac (**Methods**), overlapped with differential H3K4me1 and/or H3K4me2 sites in KS individuals compared to normal controls where the H3K4me1/me2 signal intensities were significantly reduced for the majority of them (**Figure 4B**). Genes whose super-enhancers overlap with differential H3K4me1/2 in KS include several important genes previously associated with mechanistically overlapping clinical syndromes or presentations. (**Figure 4C, Table S4**). *RAP1A* is associated with KS-like phenotypes and plays an important role in the RAS/MAPK signaling pathway.(Bögershausen et al. 2015; Tsai et al. 2018) *CHD7* is a major cause of CHARGE syndrome characterized by an overlapping clinical phenotype with KS.(Butcher et al. 2017) *PAX5* is a master regulator in B cells and is known to be a co-binding factor of *KMT2D* and implicated in the B cell development defect. (Lindsley et al. 2016) The results indicate that *KMT2D* mutations might be associated with disruption of a substantial fraction of the super-enhancers, implicated in KS pathogenesis.

**Figure 4.**
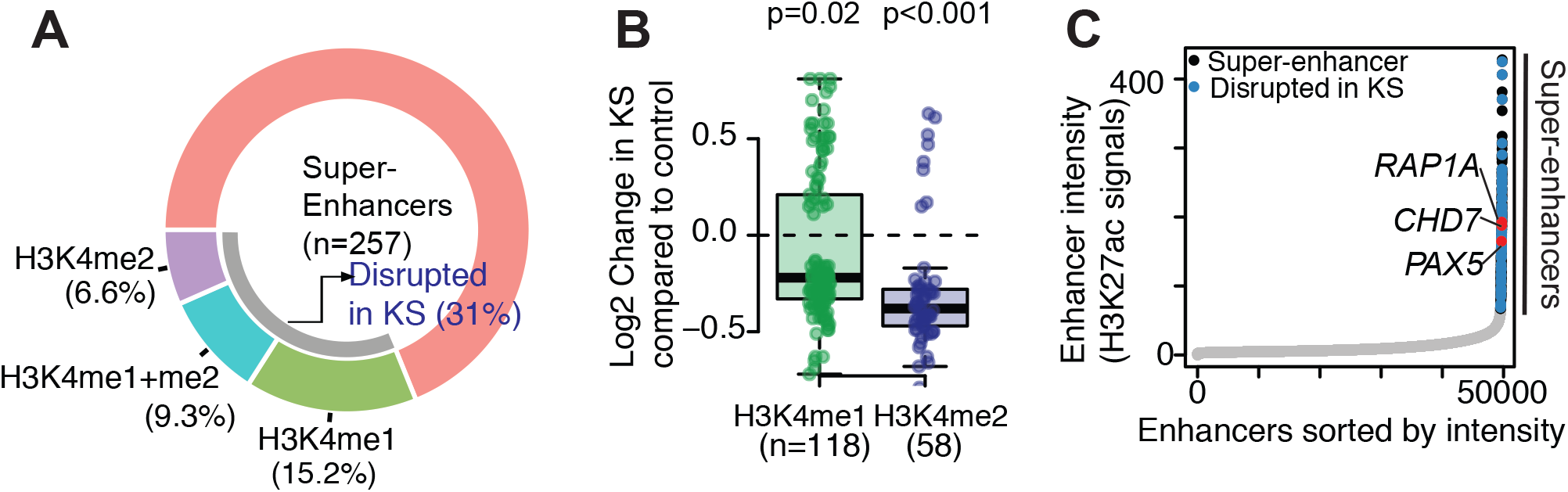
Association between altered enhancer sites in KS and super-enhancer regions. **A**. Fraction of super-enhancers overlapping with altered H3K4me1 and/or me2 peaks in KS compared to control. **B**. Fold changes of altered signals of H3K4me1 and H3K4me2 within the super-enhancers. P-value by t-test. **C**. H3K27ac signal intensities for active enhancers. Gray, non-super-enhancers; black, super-enhancers; blue, super-enhancers overlapping with altered H3K4me1/me2 in KS.

### Relationship between transcriptional changes and enhancer signal changes

To explore the expression changes downstream of changes in enhancer signals in KS, we examined the correlation between H3K4me2 and transcription changes in KS individuals compared to controls (**Methods**). For the differentially expressed genes (q < 0.1), the fold changes of H3K4me2 were modest but positively correlated with the fold changes of expression levels (Pearson’s *r* = 0.4), with approximately 70% of genes displaying changes of expression and H3K4me2 levels in the same direction as illustrated in **Figure 5A**. The downregulated genes with decreased H3K4me2 signals in KS include genes associated with immunity, linking this H3K4me2 signature to the KS phenotype. For instance, *TNNT1* was reported as one of the 20 most significantly dysregulated genes in Mll2 knockout mice and plays a role in musculoskeletal development.(Issaeva et al. 2007) *IR23R* and *MXRA8* are implicated in autoimmune disease and immune response, respectively.(Peloquin et al. 2016; R. Zhang et al. 2019) Several genes including *TUSC3* are frequently dysregulated in cancer.(Gu et al. 2016) This result indicates that changes in enhancer marks could explain expression changes for a majority of the target genes. However, additional factors besides H3K4me2 likely also contribute to the transcriptional changes in KS, given that 30% of genes do not show expression changes in the same direction as H3K4me2.

**Figure 5.**
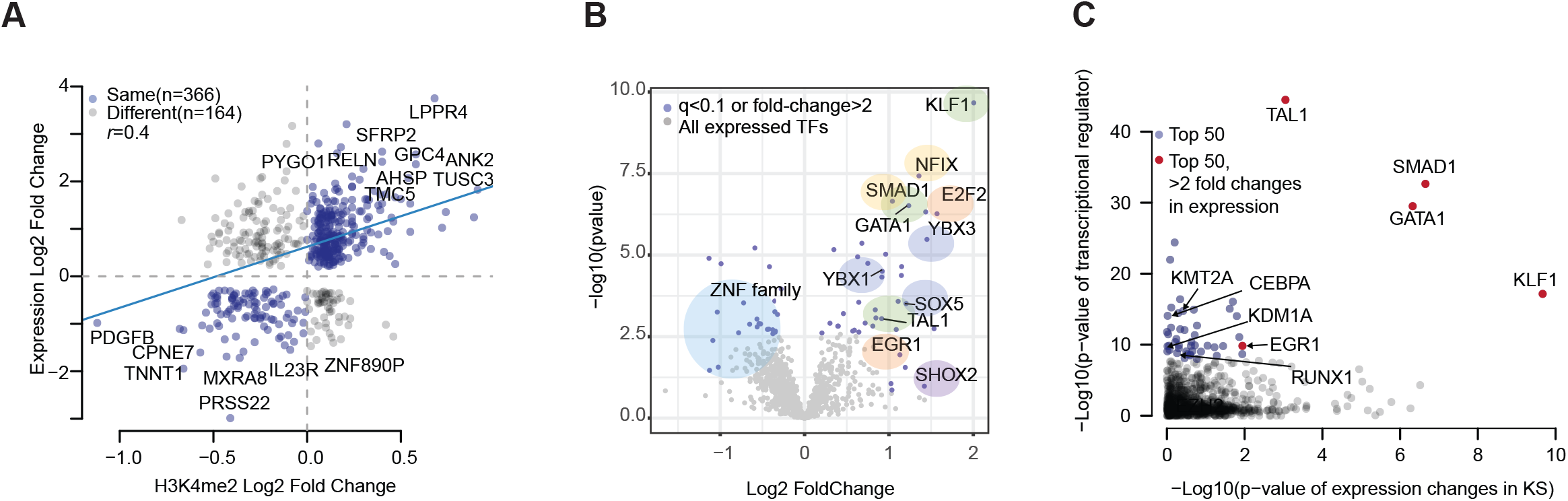
Transcriptional changes in KS. **A**. Relationship between H3K4me2 signal changes and expression changes in KS compared to control. Each dot represents each gene with significant expression changes (q = 0.1). Blue, changes in H3K4me2 and expression levels in the same direction; gray, changes in H3K4me2 and expression levels in the opposite direction. The blue line shows the regression line with a Pearson correlation efficiency of r value (r = 0.4). **B**. Volcano plot of expression changes in KS compared to control for TFs. Blue, TFs exhibiting substantial transcriptional changes in KS with q-value < 0.1 or fold changes > 2; gray, rest of TFs expressed in blood. Light blue circle, ZNF family; green, erythroid cell master regulators; yellow, hematopoietic cell regulators; orange, cell cycle genes; blue, B, Th and T cell regulation; purple, dysregulated gene in other tissues. **C**. Transcription factors regulating differentially expressed genes in KS compared to control. The y-axis represents the significance of prediction based on binding from public ChIP-seq data near the differentially expressed genes. The x-axis shows the significance of expression changes in KS for the gene. Blue, top 50 regulators underlying differentially expressed genes in KS; red, top 50 regulators underlying differentially expressed genes in KS and showing greater than 2-fold changes in expressions.

### Transcriptional regulators underlying expression changes in KS

In the previous sections, we report that (1) TFs EGR1 and E2F2 experienced substantial transcriptional changes in KS and (2) for a subset of genes, their expression changes cannot be explained by H3K4me2 changes alone. These observations prompted us to search for other TFs, which could globally affect the expression changes of the TF target genes. A total of 65 TFs exhibited noticeable expression changes in KS patients (*q*-value < 0.1 or log2 fold change > 1, **Figure 5B, Methods**) including KLF1, GATA1 and TAL1, master regulators in erythroid cells. A recent study showed the association of KLF1 in autoimmune disease.(Teruya et al. 2018) SOX5 is associated with differentiation in B cells and T helper 17 cells. Its overexpression is implicated in autoimmune disease as well.(Tanaka et al. 2014) YBX TF families are involved in T cell differentiation; NFIX and SMAD1 are associated with hematopoietic stem cell regulation.(Martynoga et al. 2013) These TF expression changes were significant compared to the expectation from the background gene expression changes (p < 10^−5^ by rank sum test, **Figure S3, Methods**).

To infer transcriptional regulators, such as TFs or chromatin regulators, underlying differential expressions in KS, we computed regulatory potential scores by integrating transcription regulator binding sites from public Chip-seq data and differentially expressed genes in KS. In brief, using about 10,000 publicly available ChIP-seq data, the regulatory potential by each transcriptional regulator was calculated for differentially expressed genes (DEGs, **Methods**). The top 50 TFs underlying DEGs in KS include TAL1, KlF1 and GATA1, master regulators in erythroid cells and SMAD1 for hematopoietic cells (**Figure 5C**). We found KMT2A (lysine-specific methyltransferase 2A; another methyltransferase for H3K4me), CEBPA (CCAAT enhancer binding protein alpha) and KDM1A (lysine-specific histone demethylase 1A) which are associated with enhancer function. However, KMT2D itself was not identified, likely due to sparsity in available ChIP-seq datasets. EGR1 and RUNX1, which are TFs predicted from the differential H3K4me2 sites in **Figure 2B**, were also included in the list. Therefore, the disrupted expression states of these TFs likely regulate downstream differential expression in KS. Although the gene expression changes may not be a direct consequence of *KMT2D* gene mutations, these results support that expression changes are a consequence of expression changes of TFs in KS.

## DISCUSSION

Although *KMT2D* is the major causal gene for KS and its primary role in chromatin states is known, the effect of haploinsufficiency of KMT2D on histone modification and subsequent transcriptional (dys)regulation has not been studied in samples from patients with KS. To examine the alterations in the chromatin landscape in KS, we profiled H3K4 methylation states from blood samples of 33 KS patients with pathogenic variants in *KMT2D* along with transcription data and systematically identified altered chromatin and expression states in KS.

Our analysis of H3K4me1/2/3 profiles from KMT2D haploinsufficient patients revealed an enhancer landscape (H3K4me1/2) in KS distinctive from unaffected controls. Interestingly, the H3K4me2 profile was more impacted compared to H3K4me1, with better separation between KS and control groups. Although both H3K4me1 and H3K4me2 are associated with enhancers, they target substantially different genomic locations as reported in previous studies.(Pekowska et al. 2010) We also observed a small number of KS-specific alterations in H3K4me3 peaks at promoter sites. However, in our dataset, the majority of H3K4me3 changes are explained by genetic background rather than *KMT2D* genotype. It is notable that the reported roles of KMT2D in literature are somewhat inconsistent, as some studies describe KMT2D as a methyltransferase specifically acting on a single mark (e.g., H3K4me3 or H3K4me1), while others describe KMT2D to act on enhancer marks (H3K4me1/2) or all forms of H3K4me.(Fahrner et al. 2019; Carosso et al. 2019; Huisman et al. 2021; Zhang et al. 2015; Ortega-Molina et al. 2015) Despite the variations and discrepancies in the literature, most studies report greater changes in H3K4me1/2 than H3K4me3. In line with the reported literature, our results show that *KMT2D* haploinsufficiency has a greater impact on H3K4me1/2, thus enhancer states.

More specifically, we found distal enhancer signals of immune-related genes or immune pathways were frequently reduced in KS, suggesting dysregulation of the associated immune-related genes/pathways, thus linking the distal enhancer states with the immunodeficiency phenotype frequently observed in individuals with KS. In addition, we observed promoter-proximal enhancer signals to be increased in genes associated with proliferation, inflammation and metabolism pathways, which is in line with previous studies in lymphoma, which showed that loss of KMT2D promotes proliferation and tumorigenesis. In contrast, results from the mouse Kmt2d model indicated that Kmt2d haploinsufficiency in neuronal cells might also affect metabolic pathways but in the opposite direction, leading to reduced proliferation. Taking together our and previous findings, a conclusion is emerging that the effect of KMT2D haploinsufficiency might be tissue- and cell-type specific.

Based on the active enhancer mark of H3K27ac profiled in GM12578, we showed that approximately one third of super-enhancer regions overlapped with altered enhancer regions in KS when compared to control, suggesting a potential impact on the regulation of super-enhancers in KS. Prior studies on the role of KMT2D in super-enhancers during tumorigenesis showed that KMT2D plays a role in maintaining super-enhancers of tumor suppressors and that the loss of KMT2D resulted in dysregulation of glycolytic genes in tumors.[reference] Our observations indicate possible disruptions of super-enhancer regulation in KS to also occur in the non-cancer context, where they are not limited to tumor suppressors or glycolytic genes. Many of the genes associated with disturbed super-enhancers in KS included genes with immune functions or B-cell differentiation. Intriguingly, super-enhancer regions of the genes implicated in KS-like phenotypes, such as *RAP1A* and *CHD7*, overlapped with altered enhancers in KS, implying a possible mechanical link with KS. Further studies are required to determine the extent of the impact on super-enhancers using H3K27ac profiled in KS individuals by haploinsufficiency of KMT2D and the functional studies on the super-enhancers of these genes.

Despite the variations in the mutational landscape of KMT2D, the resulting phenotypes in the enhancer landscape, in particular in H3K4me2, were more consistent among KS. In the current study of 33 individuals, we did not observe a clear correlation between variant types and phenotypes in chromatin or transcriptional landscape in KS. Future studies using a larger number of samples and profiles might allow us to detect the association between genomic variations in KMT2D and chromatin phenotypes.

Our RNA-seq comparison between KS and control revealed that a significant number of TFs, which are expressed in our cohort and experienced transcriptional changes, are involved in differentiation and key regulation of blood cells. A recent study in B-cells of the mouse Kmt2d model also indicated the association between Kmt2d haploinsufficiency and overall expression changes in TFs.(Luperchio et al. 2021) Although the precise estimation of expression changes of TFs in KS was limited by the heterogeneity of cell populations in our blood samples, our results support the association of transcriptional changes in TFs with dysregulation of blood cell differentiation, including known B-cell differentiation defects reported in KS phenotype(Pilarowski et al. 2020; Lindsley et al. 2016), as well as a broader range of dysregulation of the TF target genes in general.

It is pertinent to note that the data in this study represents the average signals across different cell types, making it challenging to gain insights into cell-type-specific regulation mechanisms. Profiling chromatin accessibility and transcription at a single cell level using emerging sequencing techniques in the future will allow us to better understand the regulation mechanism underlying KS in a cell-type-specific manner and to uncover novel cell types, genes and pathways that contribute to the KS etiology as well as biomarkers.

In summary, we present the first genome-wide chromatin maps of histone modifications from a large number of KS individuals with *KMT2D* mutations. Our analysis suggests unique enhancer signatures in KS with significant changes in gene regulation and regulatory pathways associated with immune, cell cycle, and proliferation as well as TF dysregulation in KS (**Figure 6**). The KS-specific chromatin changes may serve as future drug targets or biomarkers.

**Figure 6.**
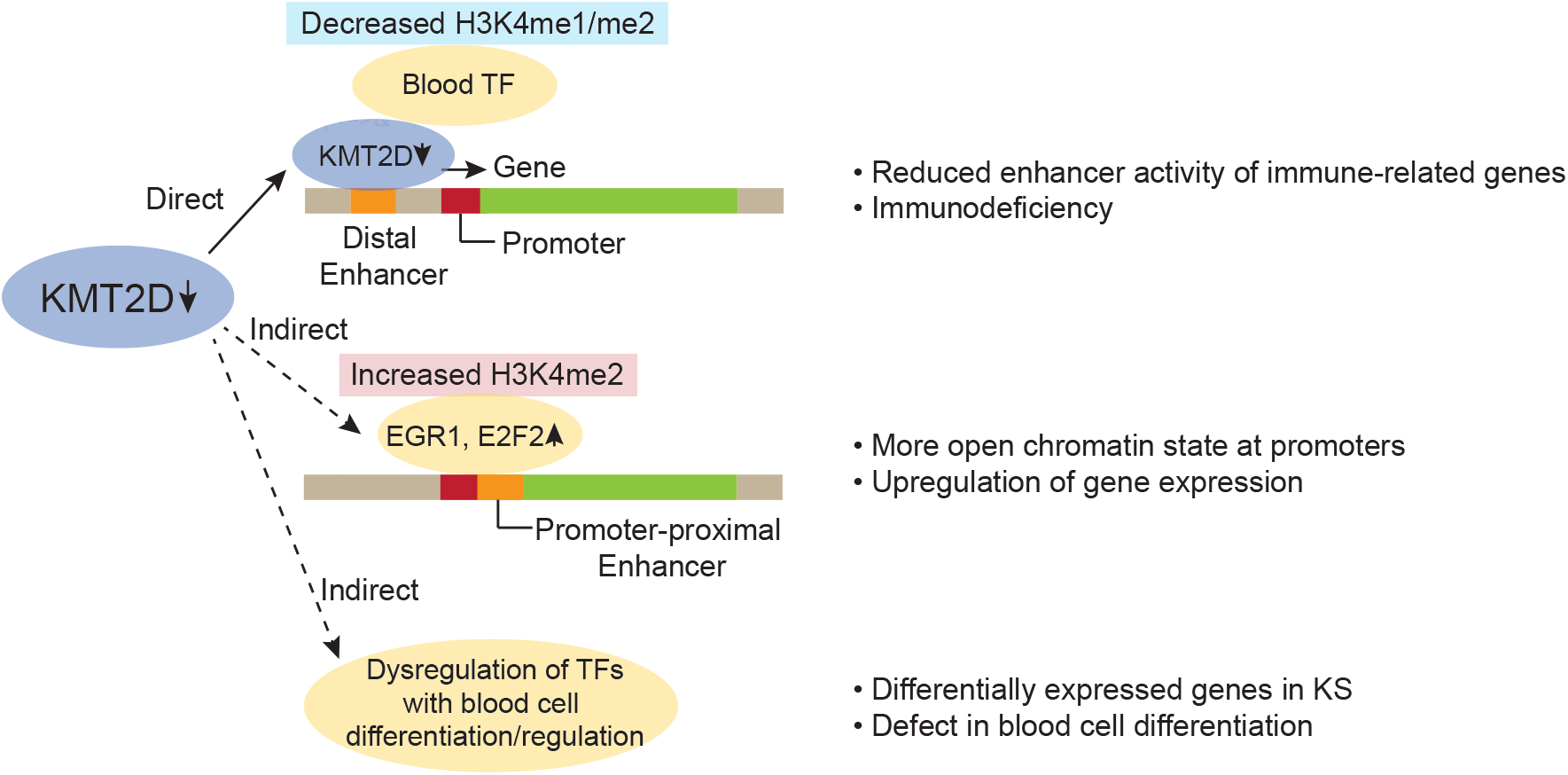
Molecular processes underlying dysregulation in KS. Schematic diagram for the underlying transcriptional regulation program in KS. Haploinsufficiency of *KMT2D* directly affects the *KMT2D* targets for enhancers/super-enhancers along with blood cell regulating TFs for immune-related genes. Upregulation of cell cycle genes, indirectly caused by haploinsufficiency of *KMT2D*, is associated with more open chromatin states near target gene promoters, resulting in the upregulation of metabolic genes. Finally, multiple dysregulated TFs affect a vast number of the target genes, which subsequently causes changes in blood cell differentiation.

## Data Availability

All data produced in the present study are available upon reasonable request to the authors

## Acknowledgments

We are particularly grateful to the patients with KS, their families and the healthy volunteers for their participation. In addition, we thank Takeda Pharmaceuticals for the financial support for the ChIP-seq data generation. Lastly, we are grateful to Tara Daly, the Roya Kabuki Program Manager at Boston Children’s Hospital for her assistance in coordinating the study visits.

## Notes

### Competing Interest Statement

The authors have declared no competing interest.

### Funding Statement

ChIP and RNA sequencing of patient samples were funded by Takeda. Takeda was not involved in data analysis and/or writing of the manuscript.

### Author Declarations

The study was approved by the Institutional Review Board of Boston Children Hospital (IRB-A00026691). All authors are affiliated with Boston Children Hospital.

