## Supplemental Figures for "Characterizing the molecular impact of *KMT2D* variants on the epigenetic and transcriptional landscapes in Kabuki Syndrome"

**Supplementary Figures**

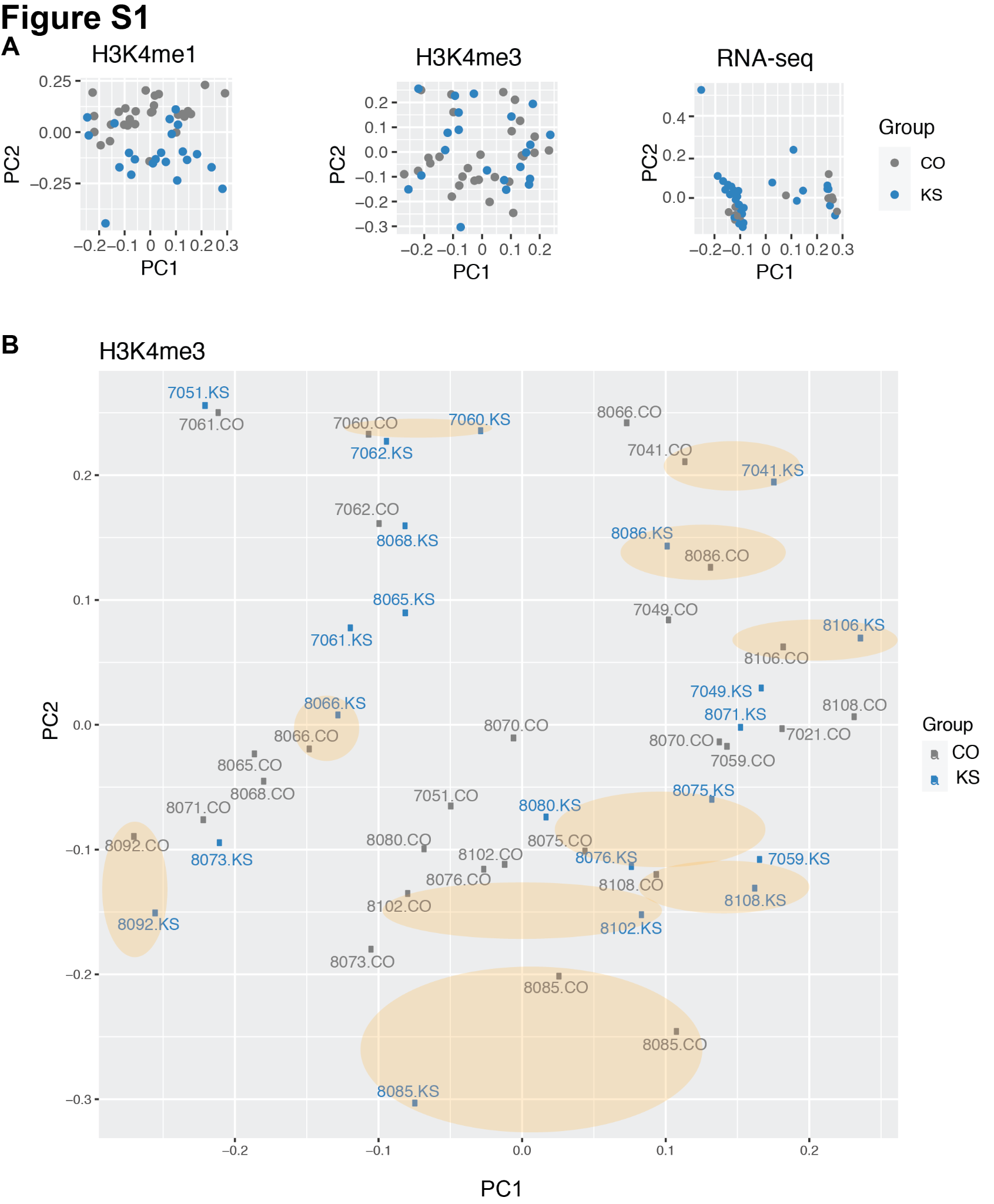

**Figure S1. Principal component analysis of H3K4me1, H3K4me3 and gene expression from KS individuals and healthy controls. A.** PCA analysis of variable H3K4me1 and H3K4me3 peaks based on ChIP-seq and RNA-seq for KS and control. Each dot represents an individual sample. Blue, KS; gray, control. **B.** PCA analysis of variable H3K4me3 peaks with family information for KS and control. Numbers represent family IDs. Circles indicate the pairs of KS patients and heathy controls from the same family (healthy siblings or parents of the KS patients). KS: Kabuki patients in blue; CO: controls in gray.

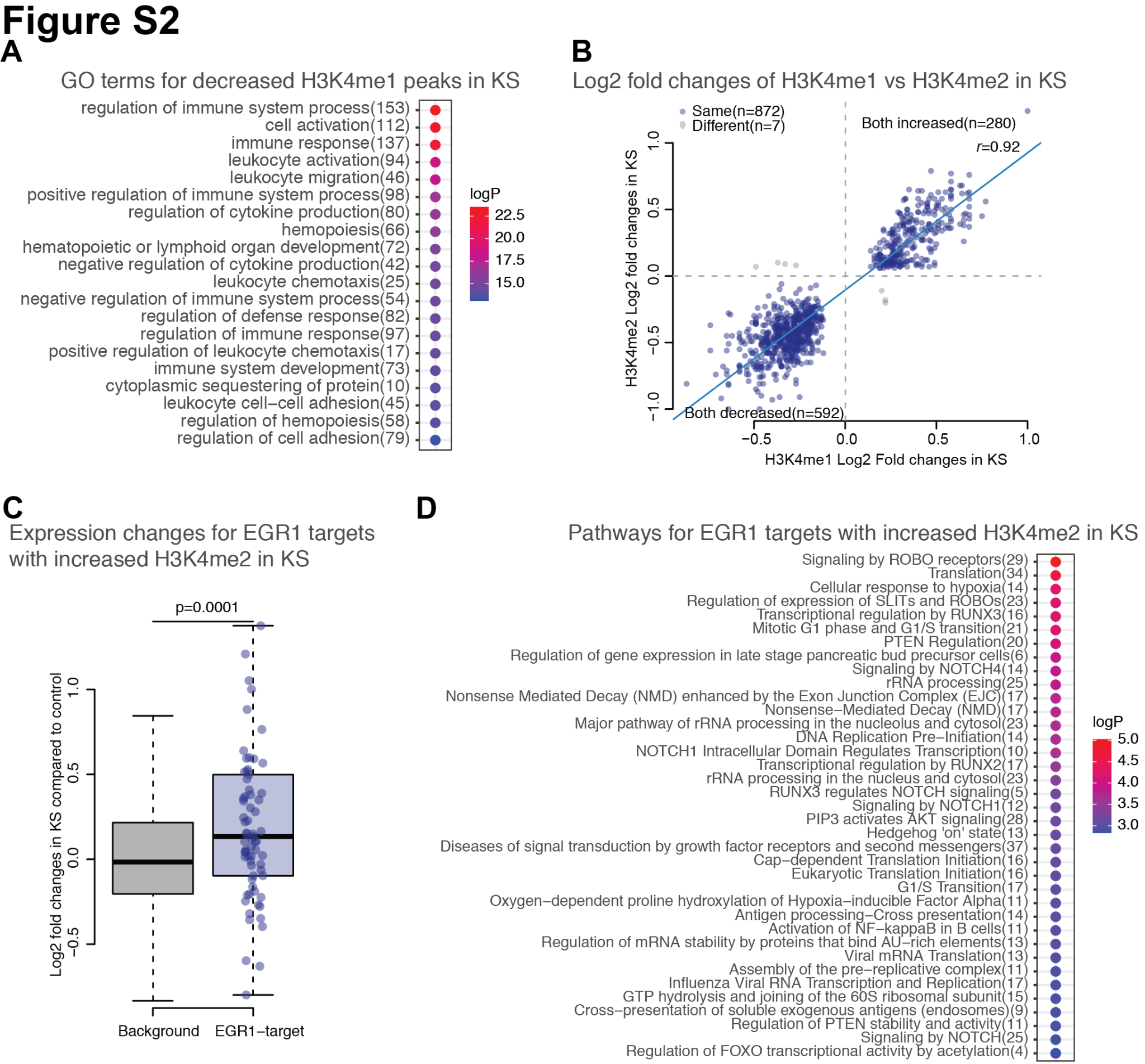

**Figure S2. Enhancer signal changes in KS individuals compared to healthy controls. A.** Top 20 gene ontology terms enriched for significantly decreased H3K4me1 peaks (q < 0.05) between KS and control. Colors represent significance of adjusted p-values of the terms. **B.** Comparison of signal changes in H3K4me2 and H3K4me1 for significantly changed H3K4me1 peak sites (q < 0.05) in KS compared to control. Blue: changes in same directions; gray: changed in different directions. **C.** Expression changes for the EGR1 target genes associated with significantly increased H3K4me2 signals (top 100) in KS compared to control. P-value by Wilcoxon Rank Sum test. Background: EGR1 target genes without H3K4me2 changes in KS. **D.** Pathways enriched for the EGR1 target with increased H3K4me2 signals in KS.

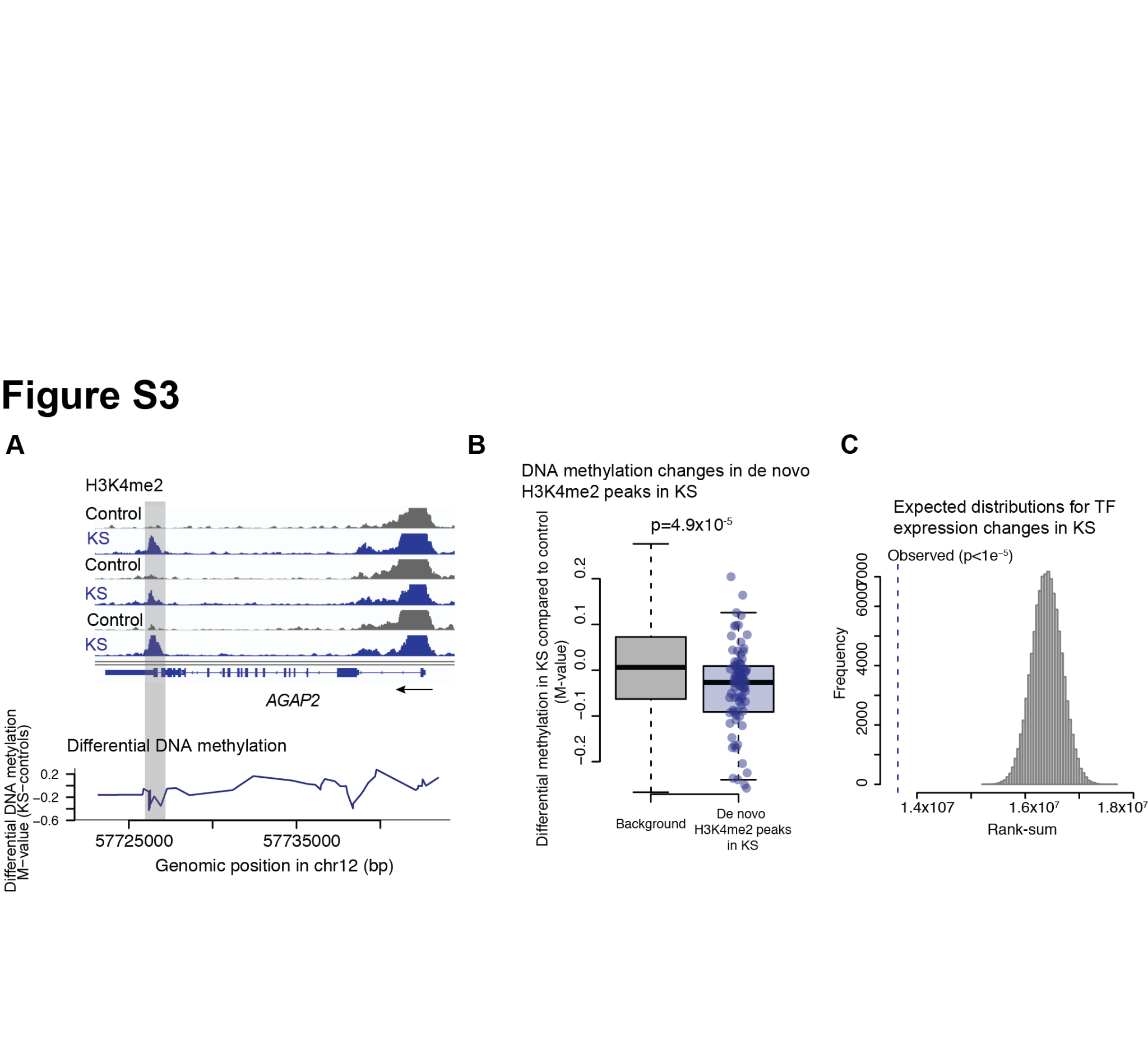

**Figure S3. DNA methylation and expression changes in KS individuals compared to healthy controls. A.** Differential DNA methylation profile of *AGAP2*. From the top, H3K4me2 profiles from KS individuals and their controls, and differential DNA methylation profile between KS and control, respectively. An arrow indicates the transcriptional direction. A gray box indicates the genomic regions overlapping with significantly increased H3K4me2 peaks and hypomethylated regions in KS compared to control. **B.** DNA methylation changes in the de novo H3K4me2 peak regions in KS compared to control. **C.** Distributions of rank-sums in expression changes for the randomly chosen genes. The dashed line indicates the observed rank-sums in expression changes for the transcription factors expressed in our KS cohort.
